# Reduced brain responsiveness to emotional stimuli with escitalopram but not psilocybin therapy for depression

**DOI:** 10.1101/2023.05.29.23290667

**Authors:** Matthew B. Wall, Lysia Demetriou, Bruna Giribaldi, Leor Roseman, Natalie Ertl, David Erritzoe, David J. Nutt, Robin L. Carhart-Harris

**Author notes:** Corresponding Author: Matthew B. Wall.

## Abstract

Psilocybin therapy is an emerging intervention for depression that may be at least as effective as standard first-line treatments i.e., Selective Serotonin Reuptake Inhibitors (SSRIs). Here we assess neural responses to emotional faces (fear, happy, and neutral) using Blood Oxygen-Level Dependent (BOLD) functional Magnetic Resonance Imaging (fMRI) in two groups with major depressive disorder: 1) a ‘psilocybin group’ that received two dosing sessions with 25mg plus six weeks of daily placebo, and 2) an ‘escitalopram group’ that received six weeks of the SSRI escitalopram, plus two dosing sessions with an inactive/placebo dose of 1mg psilocybin. Both groups had an equal amount of psychological support throughout. An emotional face fMRI paradigm was completed at baseline (pre-treatment) and at the six-week post-treatment primary endpoint (three weeks following psilocybin dosing sessions). An analysis examining the interaction between patient group (psilocybin vs. escitalopram) and time-point (pre-vs. post-treatment) showed a robust effect in a distributed network of cortical brain regions. Follow-up analyses showed that post-treatment BOLD responses to emotional faces of all types were significantly reduced in the escitalopram group, with no change, or even a slight increase, in the psilocybin group. Specific analyses of the amygdala showed a reduction of response to fear faces in the escitalopram group, but no effects for the psilocybin group. Despite large improvements in depressive symptoms in the psilocybin group, psilocybin-therapy had only a minor effect on brain responsiveness to emotional stimuli. We suggest that reduced emotional responsiveness may be a biomarker of SSRIs’ antidepressant action that is not shared by psilocybin-therapy.

## Introduction

Major Depressive Disorder (MDD) is one of the most prevalent and debilitating psychiatric disorders ^1^. Current treatment guidelines suggest psychotherapy for mild depression, with pharmacotherapy or a combination of the two treatments recommended for moderate-to-severe cases ^2^. Pharmacotherapy with Selective Serotonin Reuptake Inhibitors (SSRIs) is a popular option. However, SSRIs are only moderately effective, take four to eight weeks to show a meaningful therapeutic response, and have acceptability rates (indexed by treatment discontinuation) comparable with placebo ^3^, i.e. around 25-30% ^4^.

A phenomenon commonly referred to as ‘emotional blunting’ (i.e., a restricted range or intensity of emotional experience) has been associated with SSRI use, with one survey suggesting a prevalence close to 50% ^5^. Diminished libido and sexual functioning with SSRI treatment ^6, 7^ and a modest impact on symptoms of anhedonia in depression ^8^ are also commonly reported. One potential cause of diminished emotional responsiveness via SSRIs may be increased 5-HT activity on inhibitory postsynaptic 5-HT1A receptors in limbic and paralimbic circuitry implicated in affective and hedonic functioning ^9, 10^. Specifically, decreased amygdala responsiveness to emotional stimuli (i.e. a normalization of responses to negative stimuli) has been found in depressed patients soon after beginning a course of SSRIs ^11, 12^, and similar effects have been found in healthy subjects with short-term use of SSRIs ^13, 14^. This emotional/amygdala blunting has been hypothesized to be central to their therapeutic effect ^15, 16^.

Novel treatments for major depression are generating a great deal of interest in psychiatry ^17, 18^. Ketamine has shown potential as a rapid-acting treatment option ^19^, and promising results are being seen in small clinical trials assessing psilocybin therapy for depression ^20–22^ and depressive symptoms ^23–25^. Psilocybin is a naturally occurring ‘classic’ psychedelic which exerts its characteristic subjective effects through direct agonism of the 5-HT2A receptor ^26^. Acutely a 25mg dose of psilocybin has profound effects on spontaneous brain function ^27^ and longer-term functional brain changes have been reported with this dose of psilocybin in depressed ^28–30^ as well as in healthy individuals ^31, 32^. In clinical use, psilocybin dosing sessions can be combined with psychological therapy and support, before (‘preparation’ sessions), during the dosing sessions, and after (‘integration’ sessions)^33^.

Emotional face perception paradigms are widely used in human fMRI studies, and particularly in depression research ^34^. Using this approach in a sample of patients before and one-day after psilocybin therapy for treatment-resistant depression we previously observed an increase in BOLD responsiveness to emotional face stimuli in depressed patients^35^, which may reflect early changes in responses to treatment or ‘afterglow’ phenomena^36^, i.e., a sub-acute life in mood. Despite being directionally opposite to the BOLD-reducing action of SSRIs on emotional faces ^11, 12, 37^, this finding was predictive of longer-term treatment response ^35^.

An extension of this work identified changes in amygdala connectivity that were also associated with clinical improvements ^38^. In contrast to the emotional blunting associated with SSRIs, a sense of increased emotional connection is often reported with effective psychedelic therapy ^39^. Thus, a differential effect on emotional functioning between SSRIs and psychedelic therapy could explain their contrasting fMRI findings in emotional processing paradigms.

The aim of the present study was to directly compare the brain effects of the two treatments for the first time, in a double-blinded, randomized-controlled trial. We chose escitalopram as an active comparator for psilocybin therapy because of its high pharmacological selectivity as a 5-HT reuptake inhibitor and good antidepressant performance; in terms of both tolerability and efficacy ^3, 40^. Psilocybin therapy was carried out in a manner that was generally consistent with previous studies. The clinical procedures are detailed below and the clinical findings are fully reported elsewhere ^41^. The specific emotional facial expression paradigm employed in this study was similar to those used in previous work, e.g. ^11, 35^. The main pre-registered hypothesis for this trial was that the two treatments would have significantly different effects on brain responsiveness to emotional faces (see trial registration under identifier NCT03429075; https://clinicaltrials.gov/ct2/show/NCT03429075). This is therefore the first report of the primary study outcome.

## Methods

This study was approved by the Brent Research Ethics Committee, with additional approvals from the UK Medicines & Healthcare products Regulatory Agency, the Health Research Authority, and Imperial College London. MRI data collection, drug storage, and dispensing took place at Invicro LLC, Hammersmith Hospital, London. All subjects gave written informed consent, and the trial was conducted under the principles of Good Clinical Practice.

### Design

The full clinical study procedure is reported in ^41^. This was a phase II, double-blinded, randomized, controlled experimental medicine trial. MRI scanning was done prior to any therapeutic intervention (baseline) and six weeks and one day after the first dosing day. For the psilocybin group, they received 2 x 25mg doses of psilocybin, three weeks apart. The post-treatment MR scan occurred three weeks after the second 25mg dose. After the first of the two dosing sessions, patients in the psilocybin group were provided with a bottle of capsules containing microcrystalline cellulose (i.e. inert placebo capsules) and instructed to take one per day for the next three weeks and two per day for the final three weeks until scan visit two, which was the primary endpoint. Subjects in the escitalopram group received a 1mg dose of psilocybin on each of the two dosing visits (also three weeks apart); 1mg has negligible subjective effects and therefore served as a control procedure/placebo. These subjects were provided with encapsulated escitalopram (10mg capsules) and were instructed to take one capsule (10mg escitalopram) daily for the first three weeks, and two capsules (20mg) daily for the final three weeks. Scan visit two occurred on the day of the final capsule ingestion at the approximate time of peak plasma concentration, implying both steady state and acute presence of escitalopram for the escitalopram group.

### Clinical scales

The primary pre-registered (https://clinicaltrials.gov/ct2/show/NCT03429075) clinical outcome was the Quick Inventory of Depression Score (QIDS-SR16) ^42^. Additional measures used were the Beck Depression Inventory (BDI) ^43^, the Warwick-Edinburgh Mental Well-being Scale (WEMWBS) for assessment of well-being ^44^, the Snaith-Hamilton Pleasure Scale (SHAPS) ^45^ for assessment of anhedonia, the Laukes Emotional Intensity Scale (LEIS) ^6^ for assessment of emotional function, and the PRSexDQ ^46^ for assessment of changes to sexual function.

### Participants and Recruitment

For full details of the recruitment and screening procedures see ^41^. Participants were recruited using trial networks, social media, and other sources, via a recruitment website (https://www.imperial.ac.uk/psychedelic-research-centre). Volunteers emailed the recruitment coordinator, so all recruited participants were self-referred. There followed a multi-step screening process involving telephone and video calls, and in-person sessions with a trial psychiatrist.

Inclusion criteria were: a score of at least 17 (indicating moderate-to-severe MDD) on the HAM-D-17 assessments which were performed on the video call, confirmation of a diagnosis of depression and a satisfactory medical history (obtained from the patient’s general physician), willingness to withdraw completely from psychiatric medication (at least two weeks) and psychotherapy (at least three weeks) before starting the trial, age 18-80 years, and sufficient competence with English. The main exclusion criteria were a history of certain exclusory mental or physical health conditions (both physician-assessed), pregnancy, previous courses of escitalopram, and drug or alcohol dependence.

Thirty patients were randomized to the psilocybin arm of the study, and 29 were randomized to the escitalopram arm. However, four patients in the escitalopram arm discontinued treatment due to side effects and were therefore excluded in the present analyses. A further five subjects in the escitalopram arm did not complete the second MRI visit because of Covid-19 lockdowns in the UK in March/April 2020. Four subjects in the psilocybin arm also did not complete their second MRI scanning session also because of Covid-19 lockdowns. After the end of the trial, it was revealed that one subject in the psilocybin arm had been using cannabis regularly throughout the trial, so their data was also excluded. Consequently, there were N=21 subjects available for analysis in the escitalopram arm of the study, and N=25 in the psilocybin arm. See figure 1 for the full recruitment flow diagram.

**Figure 1.**
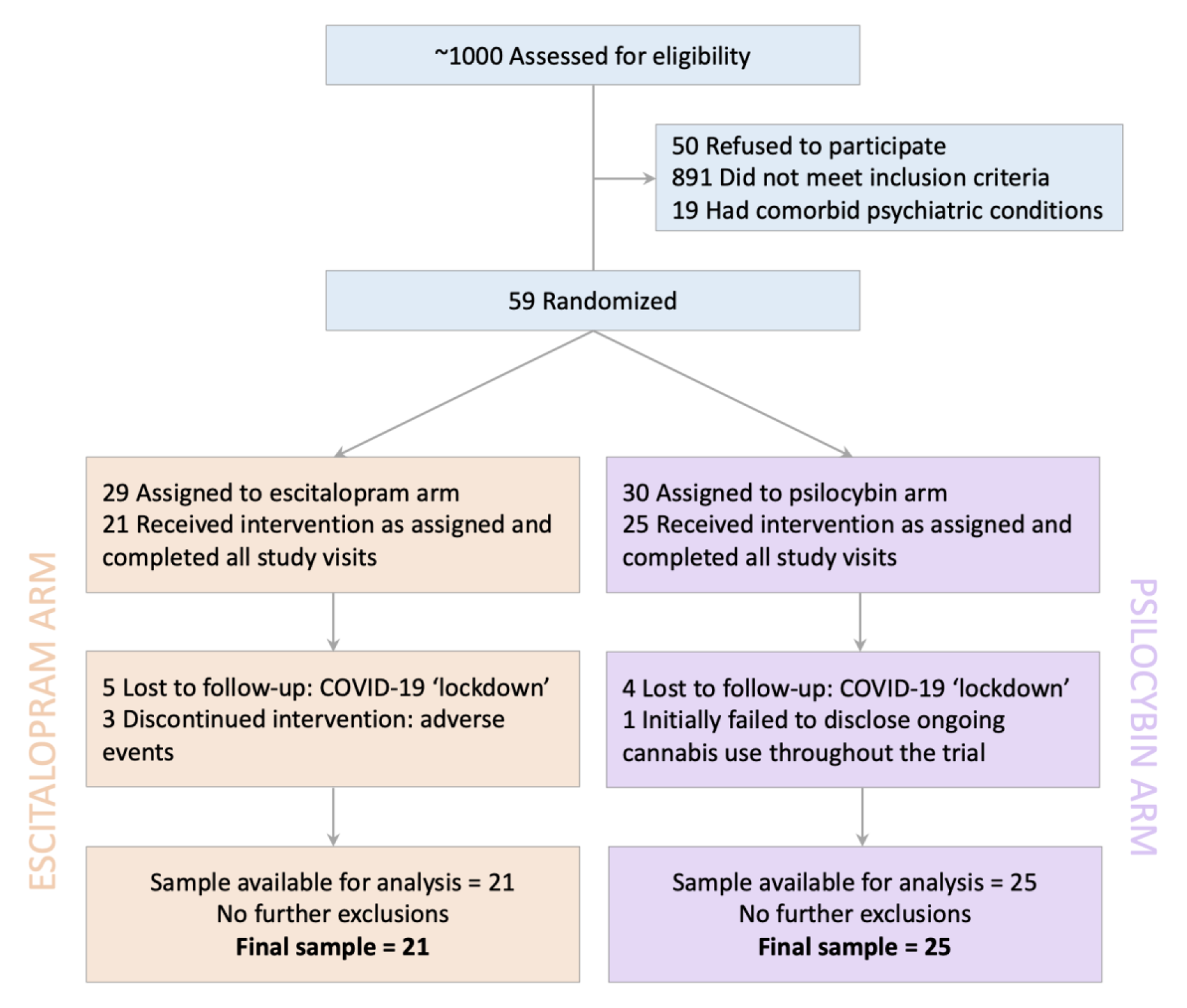
Recruitment flow diagram for the study.

### Task and image acquisition

The fMRI task involved stimuli with three facial expressions: fear, happy, and neutral. These were selected from the Karolinska Directed Emotional Faces set ^47^ and presented in blocks of five images of the same type, with each image on-screen for 3s. Rest/baseline blocks were also included, which displayed a small fixation cross at the center of the screen. There were eight repeats of each block type presented in a pre-determined pseudo-random order, and 32 blocks in total (eight each of fear, happy, neutral, and rest/baseline). The total task time was eight minutes, plus a 10s buffer period at the end of the task to ensure the last response was fully captured. Two different pseudo-random orders of block presentation were used, and subjects saw one order on the first visit and a different order on the second visit. Patients passively viewed the faces.

Data was acquired using a Siemens TIM Trio 3 Tesla MRI scanner (Siemens, Erlangen), equipped with a 32-channel phased-array head-coil. Anatomical images were acquired using the recommended parameters for MPRAGE by the ADNI-GO project ^48^: TE = 2.98ms, TR = 2300ms, 160 sagittal slices, 256×256 in-plane FOV, flip angle = 9°, 1mm isotropic voxels.

The functional (Echo-Planar-Imaging) acquisition was based on the multiband EPI WIP v012b provided by the University of Minnesota ^49–51^ using a multiband acceleration factor of 2, and a slice acceleration (GRAPPA) factor of 2 (TR=1250ms, TE=30ms, 44 slices, 3mm isotropic voxels, FOV = 192×192mm, flip angle = 70°, bandwidth = 2232Hz/pixel, 392 volumes acquired). This was based on sequences previously tested and validated by ^52^.

### Data analysis

Analysis was conducted using FSL version 5 ^53^. Processing of the anatomical data used the fsl_anat script, which performs a number of processes including inhomogeneity correction and brain extraction using BET (Brain Extraction Tool). Pre-processing of the functional data included head-motion correction, smoothing (6 mm), registration to a standard template (MNI152) using a two-step registration using the subjects’ anatomical scans, and high-pass filtering (0.01 Hz).

For the first-level analyses, a general linear model contained regressors derived from the occurrence of the stimuli in three (fear, happy, and neutral face blocks) separate regressors. These stimulus-related time-series were convolved with a standard Gamma function to model the hemodynamic response. An extended set of 24 head-motion regressors were also included as confounds, which included temporal derivatives and quadratic functions derived from the raw head-motion parameters. Modelling used FSL’s FILM (FMRIB’s Improved Linear Model) for pre-whitening and autocorrelation correction. Contrasts were computed that compared each stimulus condition with the fixation cross baseline condition, as well as a summary contrast that modelled all stimulus conditions relative to the fixation cross baseline.

To visualize treatment effects, mid-level, fixed-effects, within-subjects analyses were used to generate comparisons between pre-and post-intervention visits for each subject. Group analyses were random effects (FLAME-1) models, with statistical maps thresholded at *Z* = 2.3, and *p* < 0.05 (cluster corrected). Mean analyses including data from all subjects and all visits were generated to verify the success of the task paradigm in producing an expected pattern of brain activation, and the general acquisition and analysis procedures (see supplementary material for results). Between-subjects group-level analyses then used the results of the mid-level analyses for a comparison between the two treatment groups (psilocybin vs. escitalopram). These comparisons modelled both the difference between groups, and the difference between study visits, and therefore test an interaction effect. Data were extracted from functional clusters/maps in these group-level analyses and plotted to visualize the precise pattern of effects across the task conditions, visits, and drug treatment groups. Additionally, group-level analyses of pre-vs. post-treatment effects were also performed with an anatomical amygdala mask, derived from the Harvard-Oxford sub-cortical atlas, provided with FSL. The rationale for this analysis was based on previous work showing that amygdala connectivity and responsiveness to emotional stimuli can be specifically affected by psilocybin therapy ^35, 38^ and that it is also a region of interest in depression research ^10^. Finally, a control analysis was performed that compared the pre-therapy (baseline) scans of the two treatment groups, to examine any potential baseline differences between the two groups.

Additional exploratory analyses were conducted to examine the relationship between measures of clinical outcomes, and the BOLD activation data, using Pearson’s correlations and moderation analyses to examine the specific relationships between clinical depression outcomes, BOLD activation data, and a subjective measure of emotional function.

## Results

### Demographics

Demographic and selected clinical (baseline, pre-treatment) information for this sample are shown in table 1. There were no significant differences at baseline between the two groups on any demographic or clinical measure except for the QIDS-SR-16 score, where the escitalopram group were somewhat higher.

**Table 1.** Demographic information and selected baseline clinical scores for the sample. Pre-treatment baseline was 7-10 days before dosing day 1. All inferential statistics in the third column are non-significant (at an alpha value of *p* < 0.05) except the QIDS-SR-16 baseline score.

| Characteristic | Escitalopram<br>(N=21) | Psilocybin<br>(N=25) | Difference |
| --- | --- | --- | --- |
| **Demographic** |  |  |  |
| Age: Mean±SD (range) | 38.19±10.9 (22-60) | 42.8±11.7(21-64) | $t(44)=-1.06$,<br>$p = 0.294$ |
| Female sex: no. (%) | 6 (30) | 9 (36) | $\chi^2(1) = 0.287$,<br>$p = 0.592$ |
| White race: no. (%) | 17 (80) | 24 (96) | $\chi^2(1) = 2.67$,<br>$p = 0.102$ |
| Employment status: no. (%) |  |  |  |
| Employed | 16 (76) | 16 (64) | $\chi^2(2) = 3.55$,<br>$p = 0.471$ |
| Unemployed | 4 (19) | 7 (28) |  |
| Student | 1 (5) | 2 (8) |  |
| University education (%) | 15 (71) | 19 (76) | $\chi^2(1) = 0.12$,<br>$p = 0.725$ |
| Weekly alcohol use, UK Units:<br>Mean±SD (range) | 8.55±8.69 (0-30) | 4.8±5.8 (0-20) | $t(44)=-1.73$,<br>$p = 0.090$ |
| No previous psilocybin use: no. (%) | 15 (71) | 17 (68) | $\chi^2(1) = 0.063$,<br>$p = 0.801$ |
| Discontinued psychiatric medication<br>for trial: no. (%) | 8 (40) | 10 (40) | $\chi^2(1) = 0.00$,<br>$p = 1.0$ |
| **Clinical** |  |  |  |
| Duration of illness, years: Mean±SD<br>(range) | 15.3±11.4 (1.5-46) | 21.3±11.0(3-44) | $t(44)=-1.84$,<br>$p = 0.08$ |
| Previous psychiatric medications:<br>Mean±SD (range) | 1.95±1.63 (0-5) | 2.2±1.68 (0-6) | $t(44)=-0.505$,<br>$p = 0.616$ |
| Previous use of psychotherapy: no. (%) | 19 (90) | 23 (92) | $\chi^2(1) = 0.033$,<br>$p = 0.855$ |
| QIDS-SR-16 score at baseline:<br>Mean±SD (range) | 16.3±4.4 (6-22) | 13.9±3.4 (7-19) | $t(44)=2.128$,<br>$p = 0.039$ |
| BDI-1A score at baseline: Mean±SD<br>(range) | 28.0±6.73 (10-38) | 28.7±6.6 (16-41) | $t(44)=0.34$,<br>$p = 0.735$ |

### Clinical data

The full clinical results from the entire cohort are presented in ^41^, however, selected clinical data from the restricted cohort of n = 45 that completed the MRI scanning sessions, after exclusions, is presented here (baseline to six week timepoints). For the primary outcome measure (QIDS SR-16) a mixed-effects analysis with one between-groups factor (treatment) and one within-subjects factor (time) showed a significant main effect of treatment group (*F*[1,44] = 11.76, *p* = 0.0013) and time (*F*[6,251] = 26.36, *p* < 0.0001), but no significant interaction (figure 2A). A similar analysis was used for Beck Depression Inventory (BDI) scores, which also showed significant main effects of time (*F*[3,113] = 55.89, *p* < 0.0001), time (*F*[1,44] = 8.73, *p* = 0.005), and a significant interaction (*F*[3,127] = 6.49, *p* = 0.0004), suggesting a significantly greater decrease in BDI scores in the psilocybin group (figure 2B).

**Figure 2.**
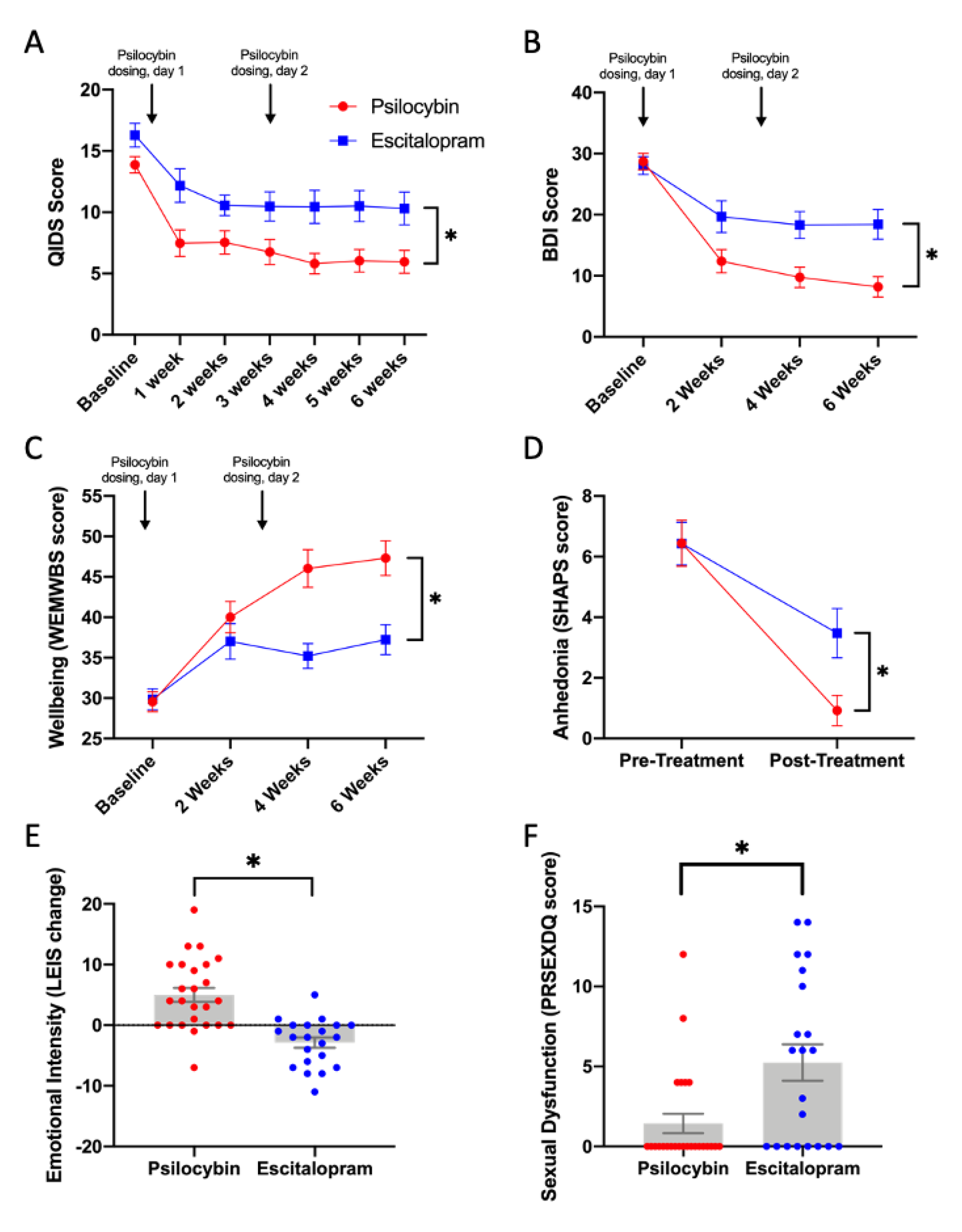
Selected clinical data from the fMRI sub-sample analyzed here. Error bars are standard errors. A: Significant interaction between the treatment groups and time on Beck Depression Inventory (BDI) scores, * *p* = 0.0002. B. Significant main effect of treatment on the Quick Inventory of Depressive Symptoms (QIDS), * *p* = 0.002. C: Significant differences between the treatment groups on the Warwick-Edinburgh Mental Well-being Scale (WEMWBS), * *p* = 0.0031. D: Significant differences between the treatment groups on the Snaith Hamilton Anhedonia Pleasure Scale (SHAPS), * *p* = 0.014. E: Significant differences between the treatment groups on change (pre-vs. post-treatment) scores on the Laukes Emotional Intensity Scale (LEIS), * *p* < 0.0001). F: Significant differences between the treatment groups on the Psychotropic-Related Sexual Dysfunction Questionnaire (PRSexDQ). * *p* = 0.0036. Note, higher scores on the PRSexDQ reflect greater sexual *dysfunction*.

The same analysis model applied to well-being (WEMWBS) data showed a significant main effect of time (*F*[2,92) = 20.93, *p* < 0.0001), a significant main effect of treatment (*F*[1,44) = 12.11, *p* = 0.0011) and a significant interaction between the two factors (*F*[3,127) = 4.97, *p* = 0.0027). These results suggest significantly greater improvement in well-being scores in the psilocybin treatment group (see figure 2C). A two-way ANOVA analysis of scores on the Snaith Hamilton Anhedonia Pleasure Scale (SHAPS) showed no main effect of treatment group (*F*[1,44) = 2.13, *p* = 0.15), but a significant effect of pre-vs. post-treatment (*F*[1,44) = 79.89, *p* < 0.0001), and a significant interaction (*F*[1,44) = 7.34, *p* = 0.0096), again suggesting greater improvement in anhedonia in the psilocybin group (figure 2D). The change (pre-vs. post-treatment) in scores on perceived emotional responsiveness or intensity (LEIS) were analysed using an unpaired *t*-test, and showed a significant difference between the groups (*t*[44] = 5.27, *p* < 0.0001), i.e., there was a relative decrease in emotional-intensity in the escitalopram group and a relative increase in the psilocybin group (figure 2E). Finally, change scores on the Psychotropic-Related Sexual Dysfunction Questionnaire (PRSexDQ) were also significantly different between the two treatment groups (*t*[44] = 3.08, *p* = 0.0036) in this restricted cohort (figure 2F).

### Imaging data

Subject head-motion was assessed and no subjects were excluded on this basis. There were also no significant differences in head-motion between groups or across study visits; see supplementary material for full details. Results of an analysis modelling the mean task activation of all subjects from each group and all scan visits showed the predicted pattern of task effects, with the emotional faces activating a broad pattern of brain regions including primary and secondary visual cortex, amygdala, thalamus, insula, and superior frontal regions (see supplementary figure S1). This pattern is entirely consistent with previous work (e.g. ^54^) and therefore validates the experimental and analysis procedures.

Results from the main analyses of treatment and drug group effects are shown in figure 3. This is a complex effect that models the interaction between the (within-subject) pre-and post-treatment factor and the (between-subjects) treatment group factor. The voxel-wise analyses found a large network of areas in which there was a decreased BOLD response post-treatment in the escitalopram group, relative to the psilocybin group. Significant clusters were evident in the dorsolateral pre-frontal cortex, supramarginal gyrus, mid-insula, and superior temporal lobe.

**Figure 3.**
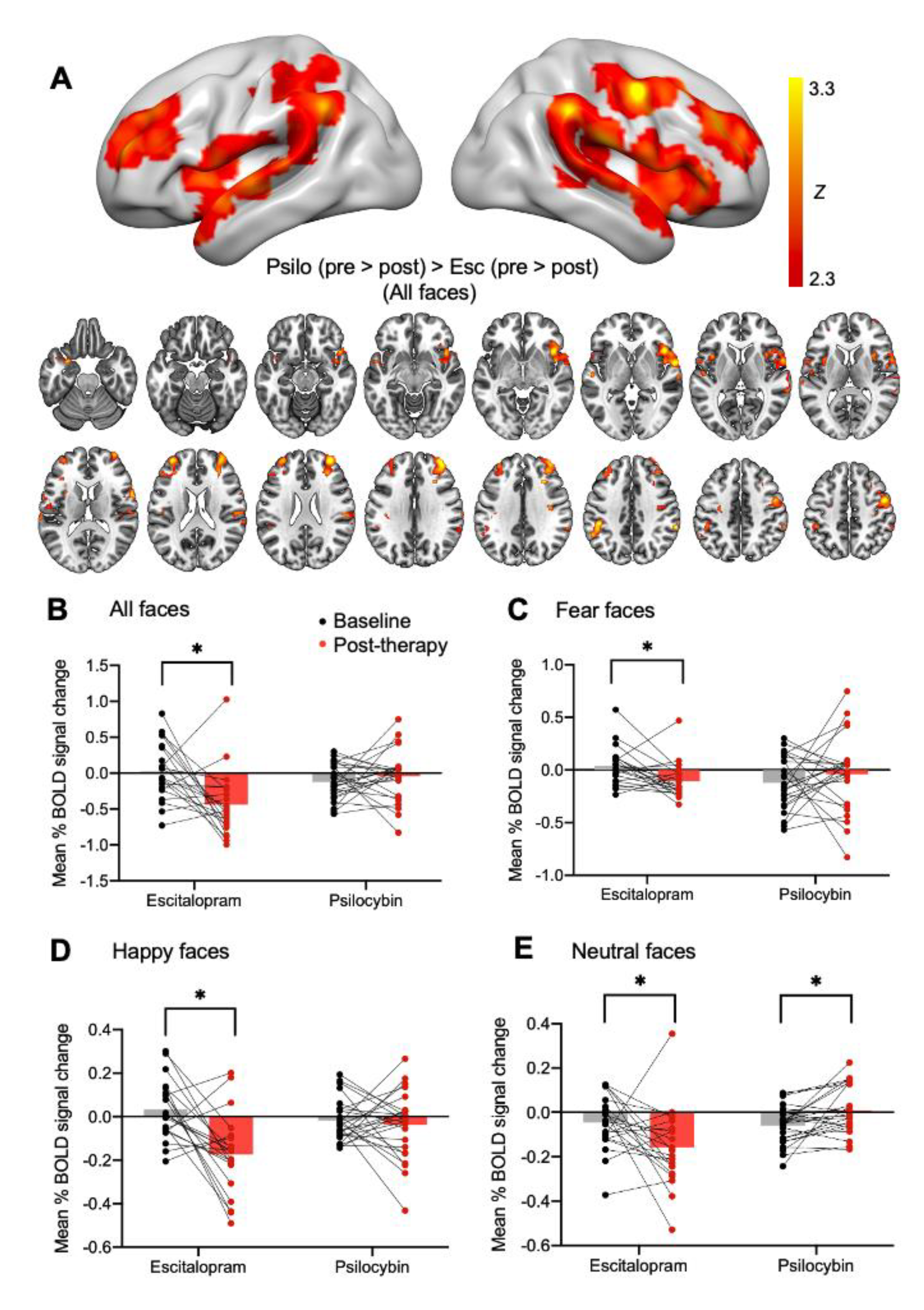
Results from the main analysis. Panel A shows results of a voxel-wise (*Z*> 2.3, *p* < 0.05, corrected for multiple comparisons) analysis of an interaction effect between the two treatment groups and study visit (psilocybin (pre-> post-treatment) > escitalopram (pre-> post-treatment), showing a relatively greater activation to faces in the post-treatment scan (relative to the pre-treatment baseline scan) for the psilocybin group, compared with the escitalopram group. Panels B-E break down this high-level effect and plot ROI data from this total set of clusters for all task conditions, groups, and study visits. Results show that the effect is driven by a decrease in response to all face types in the escitalopram group in the post-treatment visit, with minimal post-treatment change in the psilocybin group. In fact, for neutral faces there is an increased activation post-treatment with psilocybin. * = *p* < 0.05, see text for exact values.

Data was extracted from this set of clusters and plotted across all the stimulus conditions (figure 3, panels B-E). A mixed-model 2 (group) x 2 (visit) x 3 (facial expression) ANOVA showed a significant main effect of study visit (*F*[1,44] = 5.27, *p* = 0.027). There were also significant interactions between the treatment group and visit factors (*F*[1,44] = 10.83, *p* = 0.002), and the visit and facial expression factors (*F*[2,86] = 10.09, *p* < 0.001).

Follow-up comparisons revealed that in the escitalopram group there was a significant reduction in responses on the second visit (six weeks) for all three individual facial expressions: fear (*t*[20] = 2.82, *p* = 0.011), happy (*t*[20] = 3.79, *p* = 0.001), and neutral (*t*[20] = 2.25, *p* = 0.036). This effect is also significant when collapsing across all facial expressions (*t*[29] = 3.16, *p* = 0.005). All these results survive a Bonferroni-corrected *p* threshold of 0.0125, except the neutral faces result. In the psilocybin group, similar comparisons showed a significant increase in responses on the post-therapy visit for the neutral facial expressions (*t*[24] = −3.17, *p* = 0.004).This neutral faces result for the psilocybin group survived the corrected alpha threshold (*p* = 0.0125). There were no other significant effects.

Figure 4 shows the results of the analyses using a bilateral amygdala mask. These analyses compared pre-vs. post-treatment effects for the various task contrasts within each group and found a significant activation cluster in the right amygdala for the escitalopram group, with a smaller cluster in the right amygdala for the psilocybin group, both on the task contrast of fear > neutral faces (figure 3, panels A and C). There were no other significant effects on the other task contrasts conducted in these analyses using the amygdala mask. Extracting and plotting data from the relevant activation clusters (figure 3; panels B and D), and comparing baseline to post-therapy responses found that there was no change in the psilocybin group post-treatment, however, there was a reduction in responses in the escitalopram group, with a significant effect for the fear faces (*t*[20]=2.33, *p*=0.031). However, this effect does not survive a multiple-comparisons correction for the four tests conducted here.

**Figure 4.**
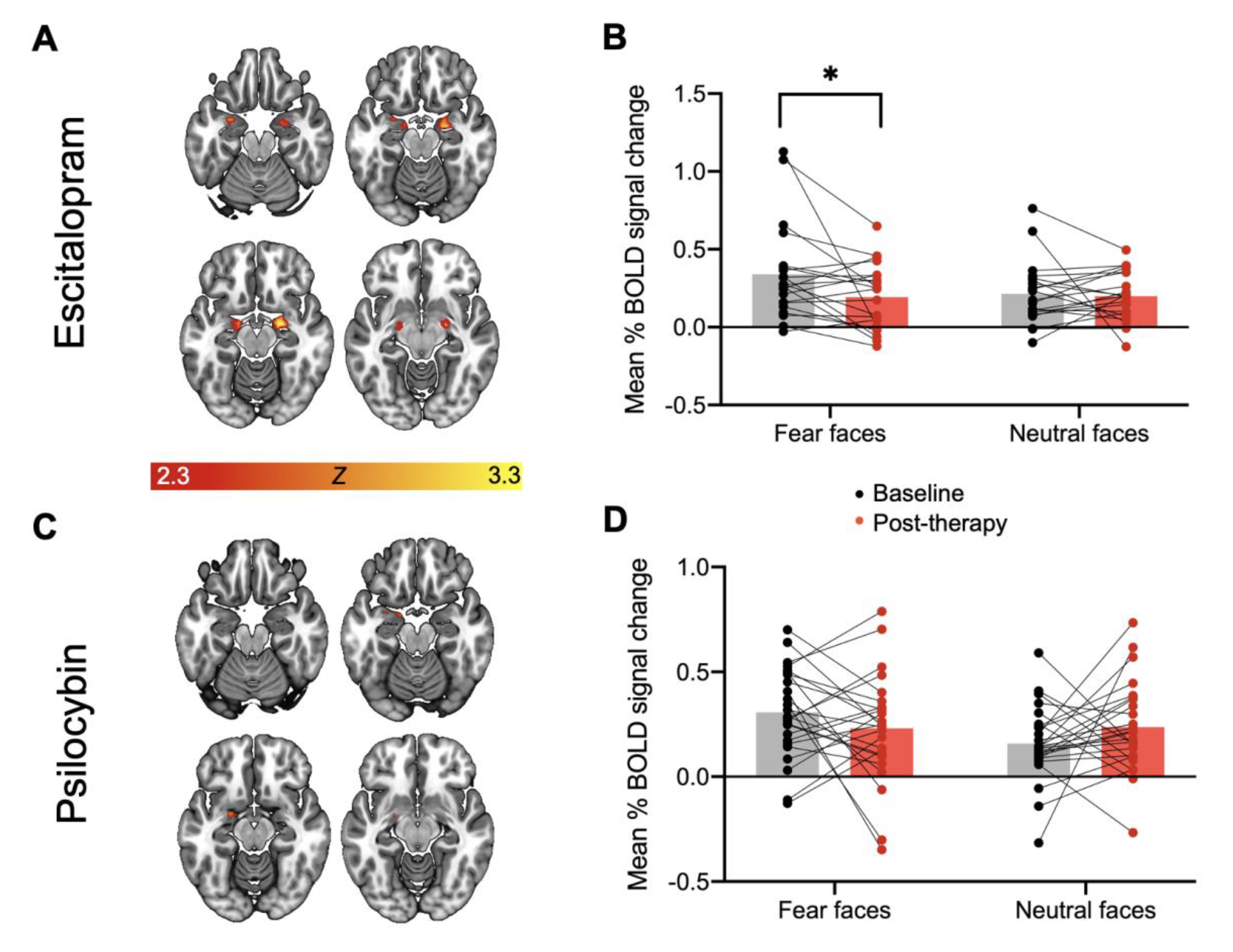
Results from analyses using an anatomical amygdala mask for the comparison of pre-vs. post-treatment for the task contrast fear > neutral. The escitalopram group showed relatively large and bilateral activation clusters for this contrast (A) whereas the psilocybin group showed a more dorsal and smaller cluster only in the left amygdala (C). ROI data from these clusters revealed a significant reduction in response to fear faces post-treatment in the escitalopram group, with no effects seen in the psilocybin group. * = *p* < 0.05.

Reassuringly, in the control analysis of the pre-treatment (baseline) scans, BOLD activations were not significantly different between the two groups on any task condition or contrast.

### Relationships between BOLD and clinical data

Exploratory analyses were conducted to examine relationships between the ROI data plotted in figure 2 and the selected clinical data presented in figure 1. Correlations were calculated within each patient group comparing the change in clinical scores (i.e., score at baseline, subtracted from the 6-week follow-up) with the change in BOLD activity (visit 1 subtracted from visit 2) from the regions identified in figure 2. There were no significant correlations present in these analyses. Results can be found in the supplementary material.

Additional exploratory work used two moderation analyses to test whether the change in brain activity (average of all faces) could predict the main clinical outcome (change in QIDS scores from baseline to six weeks), and whether this was moderated by emotional function (change in score on the LEIS). In the psilocybin group, the LEIS alone was significantly predictive of QIDS outcomes (Z = −2.24, *p* = 0.025), but brain activity alone was not (Z = - 0.07, *p* = 0.944), and neither was the interaction between the predictor and moderator (Z = - 0.91, *p* = 0.363). For the escitalopram group, the change in brain activity was predictive of BDI outcomes (Z = 1.97, *p* = 0.048), and while the LEIS alone was not (Z = 0.36, *p* = 0.721); the interaction (i.e., a moderation of the predictive power of brain activity via LEIS) was strongly significant (Z = 3.14, *p* = 0.002).

In moderation analyses, the interaction term is the most instructive result, and a significant result denotes that the relationship between the predictor and the dependent variable is stronger or weaker, given varying levels of the moderator. In this case, since all three variables are negative (i.e. relative decreases in brain activity, decreases in depression, and decreases in emotional function) it implies that - in the escitalopram group - the relationship between brain function and the clinical outcome (QIDS) is stronger when patients have greater levels of emotional blunting (i.e. lower scores on the LEIS). LEIS scores were relatively increased in the psilocybin group, meaning that the significant direct effect of the LEIS on clinical outcome (Z = −2.24) implies that a higher level of emotional function post-psilocybin-therapy relates to a greater post-treatment reduction in depression scores (i.e. greater clinical effect) after the psilocybin-therapy. These analyses were also repeated with the BDI as the dependent variable and showed a similar pattern of results; see supplementary material.

**Figure 5.**
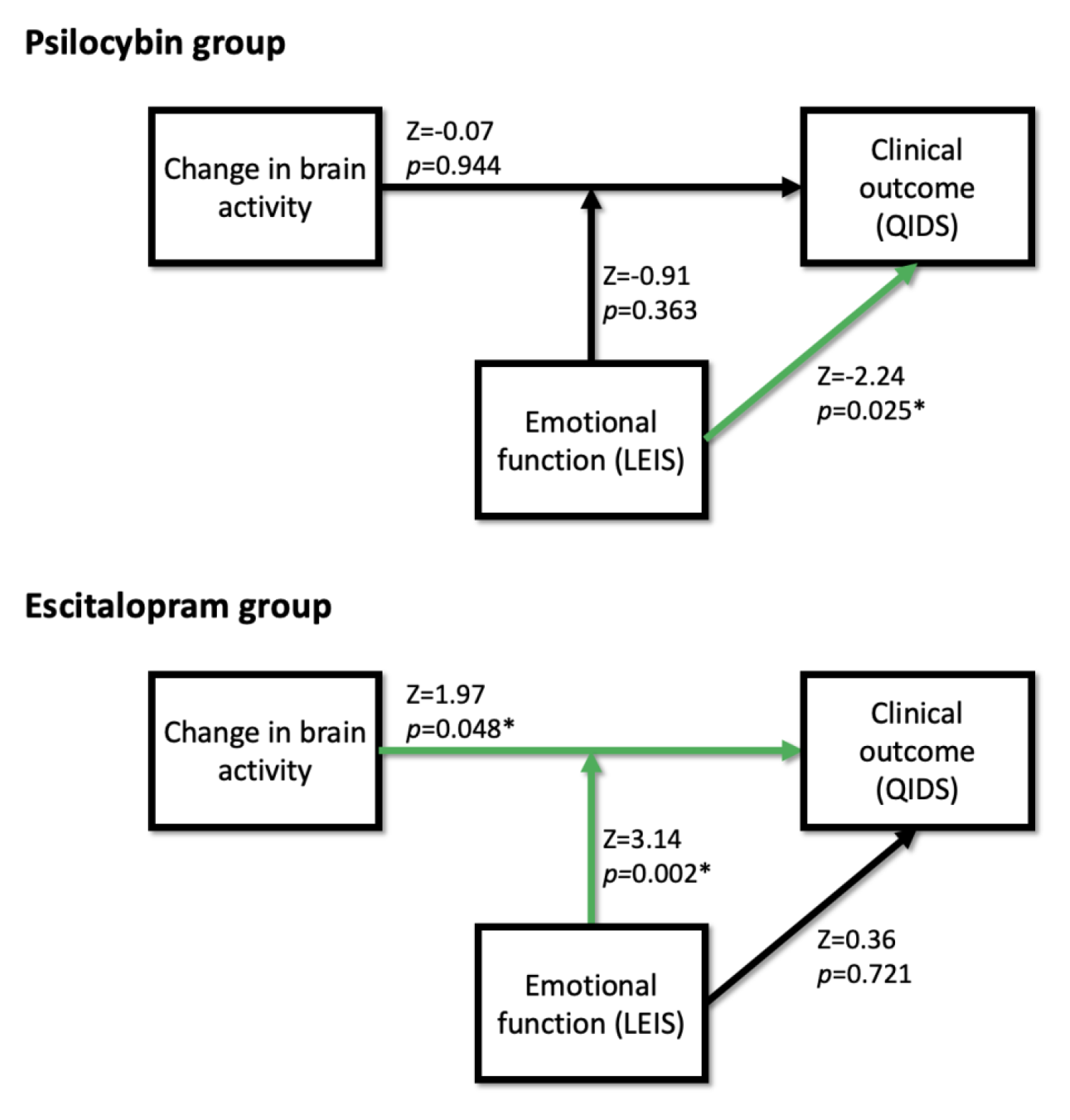
Results of exploratory moderation analyses examining the effects of the change in brain activity on the primary clinical outcome (QIDS) and its moderation by a measure of emotional function (LEIS). In the psilocybin group (top) emotional function has a significant effect on clinical outcomes, but there is no significant effect of brain activity and no interaction effect. In the escitalopram group (bottom) the change in brain activity has a robust effect on clinical outcome, and emotional function also has a strong moderating effect, supporting mechanistic assumptions about the action of the SSRI. Significant effects are denoted by a green arrow and *.

## Discussion

Consistent with our primary hypothesis for this comparative mechanisms trial, we found significant differences in brain responsiveness to emotional stimuli after escitalopram compared with psilocybin for depression. The separate examination of treatment effects revealed that the between-group differences were largely driven by decreases in responsiveness post-treatment in the escitalopram group. Null, or opposite (i.e., slightly increased activation) effects were seen in the psilocybin group, depending on the exact face type i.e., increased activations post-treatment to neutral faces was seen in the whole brain analysis. Results were generally consistent whether examining whole brain activations or focusing specifically on the amygdala. Exploratory moderation analyses suggested that outcomes were better predicted by the change in brain activity, moderated by changes in emotional function, in the escitalopram group, whereas clinical outcomes were better predicted by changes in emotional function in the psilocybin group.

This study’s findings lend support to the view that psilocybin-therapy and SSRIs have distinct therapeutic mechanisms of action ^9^. While both drugs act on the 5-HT system, SSRIs increase synaptic concentrations of 5-HT by blocking its reuptake, whereas psilocybin (through its main active metabolite, psilocin) acts as a direct agonist on certain 5-HT receptors, with a key action on the 5-HT2A receptor subtype ^26, 55^. Increased synaptic 5-HT via SSRIs should affect all available 5-HT receptors but there is some evidence to suggest that the activation of inhibitory postsynaptic 5-HT1A receptors, which are heavily expressed in limbic regions ^56^ and implicated in emotional ^57^ and sexual functioning ^58^, play an important role in the action of SSRIs. The four to six week lag in antidepressant effects with SSRIs is thought to be due to a gradual down-regulation of inhibitory 5-HT1A pre-synaptic auto-receptors ^59^, allowing a slow disinhibition of serotonergic efflux onto postsynaptic targets. Although speculative, activating inhibitory postsynaptic 5-HT1A receptors in limbic regions may dampen their responsiveness and account for the emotional blunting and loss of sexual function described by some patients receiving treatment with SSRIs ^7^. The moderation analyses provided here suggest that this reduction in brain responsiveness to emotion with SSRIs may be an important factor in their therapeutic action.

The results of the whole-brain between-groups contrast revealed a pattern of reduced responsiveness under escitalopram in regions including the DLPFC, temporo-parietal junction (TPJ), supramarginal gyrus, and secondary somatosensory cortex. These are largely high-level transmodal regions implicated in a broad range of processes, including social-cognitive functions. They are not necessarily regions most commonly identified in emotional face perception ^54^, however, a recent meta-analysis ^60^ highlighted a similar set of regions involved in empathy for physical pain, emotional situations, and emotional faces. A further recent meta-analysis of the neural effects of antidepressants as assessed by emotional response paradigms has also identified effects in a wider set of cortical regions than just the amygdala ^10^. Although speculative, the relative preservation of responsiveness in these high-level cortical regions after psilocybin therapy may relate to its positive modulation of socio-emotional functioning ^61^, an assumption that might also align with the greater responsiveness to neutral faces post psilocybin therapy.

Psilocybin therapy is emerging as a potential paradigm-shifting^62, 63^ treatment for depression and other commonly comorbid disorders^64^; paradigm-shifting because it is arguably the first truly effective drug-assisted psychotherapy^41^. This may be because psilocybin enhances affective psychotherapeutic processes in a different way to other compounds that have been combined with psychotherapy ^65^ including SSRIs^66^. Psilocybin therapy appears to have an efficacy at least comparable to established treatments^20, 22, 41^, with treatment response rates of approximately 70% in three recent trials in MDD^21, 22, 67^, and has a favorable side-effect and patient acceptance profile. The present finding of a robust difference in the effects of an SSRI and psilocybin therapy on brain responsiveness to emotional stimuli are consistent with previous assumptions about their differential therapeutic actions^9^, as well as this trial’s pre-registered primary mechanistic hypothesis (https://clinicaltrials.gov/ct2/show/NCT03429075).

A generalized emotional blunting is often associated with SSRIs^5^, whereas greater acceptance of emotions and a multi-faceted sense of reconnection^39, 68^ is commonplace with psilocybin-therapy. The differing pattern of effects in the exploratory moderation analyses presented here also support this perspective. In the psilocybin group, the (relatively increased) emotional function was shown to have a direct positive effect on clinical outcome, such that higher levels of emotional function were associated with greater improvements in clinical symptoms. In the escitalopram group however, the effect of changes in emotional function was a robust moderator of the effect of brain responses on the clinical outcome, but in an opposite direction. The relationship between brain function (decreased responsiveness to faces) and clinical effects (decreased symptom severity) was strongest for those patients who reported the largest muting of the intensity of their emotions (LEIS). As others have recently noted^9, 16^ this emotional blunting associated with SSRIs may be a key factor in their therapeutic efficacy, and the current findings support this perspective.

The importance of the 5-HT2A receptor for the action of psychedelics is well established^26^, as demonstrated for example, via a strong and selective affinity-to-potency relationship ^69^, antagonist pre-treatment blocking the psychedelic effects^70^, and most recently, subjective psychedelic effects correlating with degree of 5-HT2A receptor occupation in the human brain^71^. 5-HT2A receptors are highly expressed throughout the human cortex^56, 72^, but particularly in high-level associative/transmodal regions that undergo marked expansion throughout brain development^73^ and have expanded most evolutionarily^74, 75^. A growing body of evidence has linked 5-HT2A receptor signaling with increases in a variety of markers of neuroplasticity^76, 77^. The most recent model of psychedelic therapy’s mechanism of action emphasizes a role for enhanced 5-HT2AR-induced cortical plasticity; opening a window for healthy psychological change, guided by concomitant psychotherapy^64, 78^. In contrast to the effects of SSRIs, suppressed emotional or limbic responsiveness is not a feature of the psychedelic therapy model. Indeed, acute emotional release during ‘the trip’^79^ and then progress towards greater emotional acceptance and ‘reconnection’^39^ are thought to be important components of the psychedelic therapy model. Together with the present results, replicated findings of increased whole brain functional integrity after psilocybin therapy for depression ^30^, as well as increased dynamic flexibility ^29^, imply a different antidepressant action for psilocybin relative to SSRIs.

Regarding this study’s design, it should be noted that the final post-treatment scan occurred hours after the final dose for those in the escitalopram group (coinciding with peak plasma concentration of the drug) but three weeks after the second of two 25mg psilocybin sessions for the psilocybin group. The smaller changes in post-treatment brain functioning in the psilocybin group may be due to the duration since dosing and a different result may have been found had we scanned closer to the last psilocybin dosing session, as we did in previous work, where we showed a relative increases in amygdala responses one-day post-dosing ^28^. We believe that the present study’s design can be justified however, as the intention was to capture the enduring antidepressant effects of psilocybin-therapy, and these were robust at the 6-week endpoint (3 weeks after the final psilocybin dosing session)^41^. Our current findings can be seen as consistent with previous work in which healthy volunteers received a single-dose of psilocybin and reduced amygdala responses were observed at a one-week follow-up, with responses returning to baseline after one month ^80^. Interestingly, a distinct metric (whole-brain network modularity applied to spontaneous brain activity) has been found to reliably index the antidepressant action of psilocybin-therapy, but not escitalopram^30^. The implication is that (unlike for SSRIs^11^) the emotional face paradigm may not be an especially sensitive marker of the antidepressant action of psilocybin-therapy (on this time-scale), whereas other brain indices may be more sensitive to its specific action.

Some previous work has suggested that SSRIs and related antidepressant drugs have a selective action on the processing of negatively valenced emotional stimuli ^12^. We did not replicate this result in the whole-brain analysis but we did see suggestions of less suppression of amygdala responses to happy and neutral faces with escitalopram relative to its effect on fearful faces. However, consistent with observations from previous work ^81, 82^, faces of a negative valence evoked the largest amygdala responses in this study (figure 3). In alignment with statistical principles, one could suggest that a drug’s modulatory action would be greatest in the domain of functioning that is especially sensitive under baseline conditions, i.e. in this case, the negatively-valenced domain. It seems plausible therefore that this could account for SSRI’s apparent biased action on negatively-valenced emotional stimuli as seen in previous studies ^12^.

In conclusion, consistent with the primary hypothesis for this trial, fMRI analyses revealed a reduced brain responsiveness to emotional face stimuli after six weeks of daily escitalopram but not three weeks after the second of two 25mg dosing sessions with psilocybin-therapy for major depressive disorder. The escitalopram data is consistent with current theories on the therapeutic action of SSRIs ^15^ and the comparison with psilocybin-therapy highlights the different brain actions of these two different treatment modalities.

## Data Availability

All data produced in the present study are available upon reasonable request to the authors

## Supplementary material

### Head movement

Overall head-motion characteristics for all subjects were excellent. Assessment of subject head-motion during the scans was performed using the displacement time-series produced during preprocessing of the functional data. These were collated, and means were calculated across the time-series and across the six (three translations, three rotations) displacement parameters. Maximum displacement values were also assessed.

Only one subject showed a maximum displacement value greater than one voxel dimension (3mm) on a single parameter, for one study visit. Inspection of this subject’s time series showed a single large (∼3mm) shift in position part-way through the scan. This subject’s mean displacement (0.088mm) was less than 0.1mm, and was comparable to other subjects in the sample. It was therefore judged to be acceptable, and the subject was included in the analyses. Mean displacement across all subjects, scan sessions, and parameters was - 0. 00058mm (SD = 0.027mm).

A 2 (treatment group) by 2 (study visit) mixed-model ANOVA was conducted to examine any systematic differences in head-motion across these factors. There was no main effect of treatment group (*F*[1,44] = 1.84, *p* = 0.182) and no main effect of study visit (*F*[1,44] = 0.11, *p* = 0.740. There was also no interaction effect (*F*[1,44] = 0.077, *p* = 0.782).

### Group-level task results

**Figure S1.**
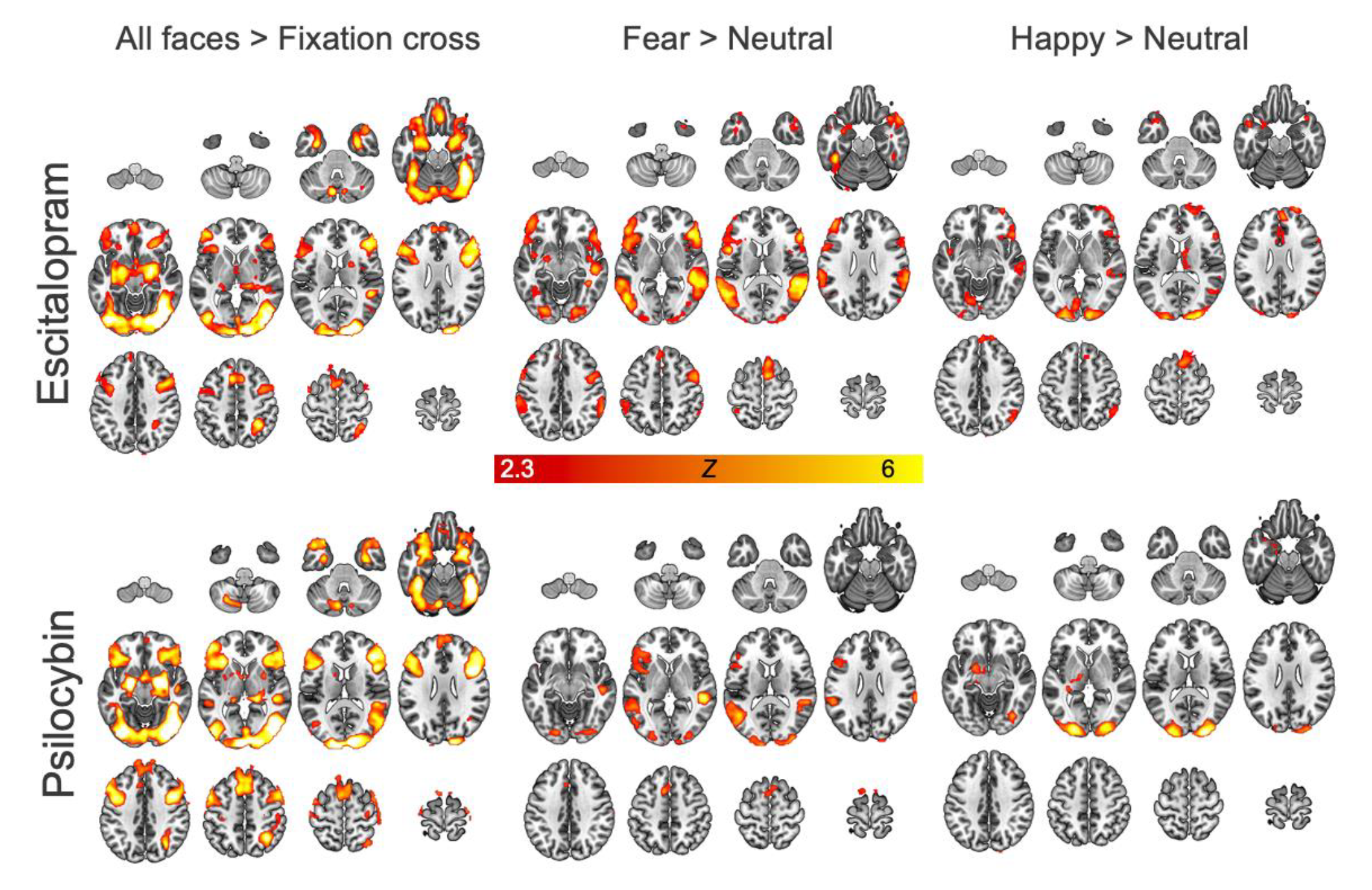
Results from the validation analyses of mean task effects in each group.

### Correlation results

**Table S1.**
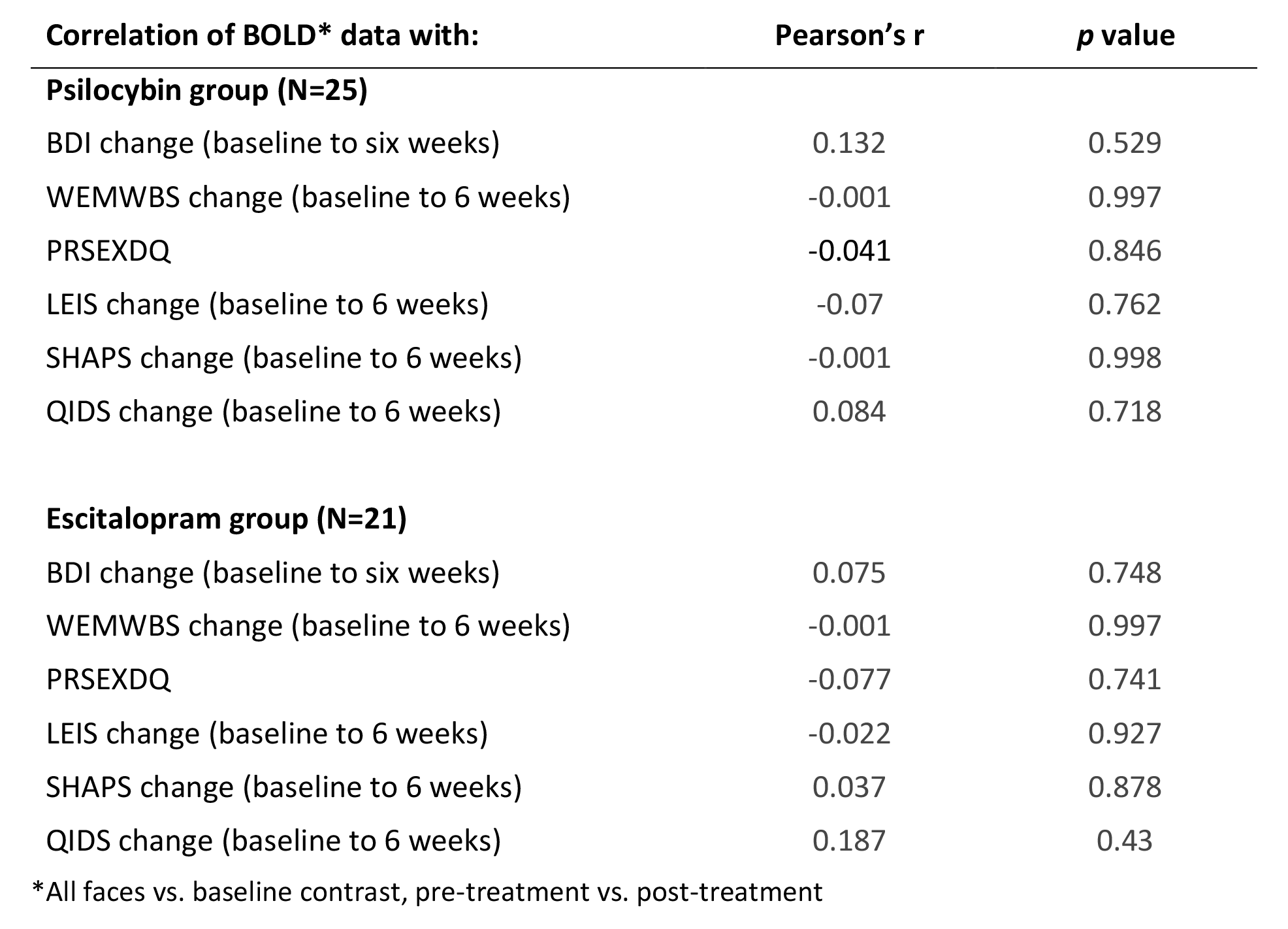
Correlation analyses between a summary measure of brain activity and key clinical measures. All analyses showed non-significant/null results.

### Moderation Analyses with BDI as the dependent variable

The moderation analyses were repeated with BDI change scores (baseline to six-week follow-up) as the dependent variable. For the psilocybin group, this produced the same pattern of results as the analysis with QIDS scores in the main text. There was a significant (*Z* = −3.02, *p* = 0.003) effect of the LEIS measure on BDI scores, but no effect of brain activity (*Z* = −0.29, *p* = 0.774) and no interaction effect (*Z* = 1.32, *p* = 0.187). In the escitalopram group there were no main effects of either brain activity (*Z* = 1.49, *p* = 0.137) or LEIS (*Z* = −0.59, *p* = 0.551) on BDI change scores, but a significant interaction effect (*Z* = 2.38, *p* = 0.018).

